# Obesogenic Memory Beyond the Body: Integrating Biological and Sociocultural Dimensions

**DOI:** 10.64898/2026.02.17.26346482

**Authors:** Varvara Borisova, Jan Gojda, Tereza Stöckelová

## Abstract

**Introduction:** Mechanistic research has shown that prior obesity induces durable transcriptomic and epigenetic reprogramming in adipose tissue that persists after weight loss and predisposes individuals to weight regain. This phenomenon, termed obesogenic memory (OM), is currently conceptualized primarily as a molecular process. We propose extending OM beyond adipose tissue biology to include interacting biological and sociocultural processes through which past exposures shape present physiological regulation and health-related behavior.

**Methods:** In-depth qualitative interviews were conducted with individuals living with obesity (n=31) and with healthcare professionals (n=18). The data were analyzed abductively to examine participants’ lived experiences of obesogenesis.

**Results:** We developed a three-phase model of OM comprising memorizing, remembering, and rescribing. The memorizing phase describes the initial acquisition and encoding of biological and sociocultural obesogenic influences. The remembering phase captures the persistence of these influences, contributing to long-term obesity maintenance. The rescribing phase refers to processes through which obesogenic influences may be attenuated or reversed, creating conditions for sustainable health behavior change.

**Conclusion:** Extending OM to include sociocultural dimensions provides a more comprehensive understanding of obesity persistence. This integrative framework identifies multilevel targets for obesity prevention and treatment that acknowledge past exposures while supporting resilience and long-term weight management.

## Introduction

Extensive research has demonstrated that obesity emerges through shared genetics, family environments, and household practices [1, 2], as well as through broader socioeconomic, political, environmental, and material conditions [3, 4]. These structural determinants have prompted calls for systemic interventions, including reforms in food policy and environmental regulation. However, such approaches have only been partially translated into public health measures, and obesity continues to be treated predominantly at the level of individual bodies, through biomedical interventions.

This individualization is likely to intensify with the growing availability of highly effective anti-obesity medications, particularly glucagon-like peptide-1 receptor agonists (GLP-1RA), which enable substantial weight loss at the individual level [5]. While clinically valuable, these pharmacological advances risk reinforcing a conceptual narrowing of obesity as a problem to be managed within individual bodies, rather than as a condition produced and sustained by collective social and material conditions. In this context, bodies may increasingly function as sites of downstream intervention, while upstream drivers of obesity—such as food systems, marketing practices, and socioeconomic inequality—remain insufficiently addressed.

In metabolic medicine, the persistence of metabolic states beyond their original conditions has long been recognized. The concept of metabolic memory, developed in diabetes research, demonstrated that early exposure to poor glycemic control leaves lasting biological imprints. The Diabetes Control and Complications Trial showed that individuals exposed to suboptimal glucose regulation remain at elevated risk for complications even after later improvements [6]. Similarly, the thrifty phenotype hypothesis proposed that undernutrition during critical developmental periods permanently alters metabolism, which increases susceptibility to metabolic disease in adulthood [7]. Together, these frameworks established that early exposure produces durable physiological programming—a form of biological memory shaped by early environments.

This insight was later extended beyond individual development to intergenerational processes. Metabolic alterations can be transmitted through epigenetic modifications of the germline, including DNA methylation, histone modification, and non-coding RNAs [8]. Some epigenetic marks escape reprogramming and persist across generations, influencing appetite regulation, adipogenesis, and energy expenditure in offspring [9]. These findings position metabolic memory as temporally deep and biologically embedded.

Human metabolism is also shaped by more-than-human ecologies. Coevolved microbial communities encode past dietary patterns and inflammatory states, functioning as a form of metabolic memory that influences appetite regulation, energy harvest, and metabolic risk [10, 11]. Modern lifestyles— antibiotic use, urbanization, formula feeding, and low-fiber diets—have reduced microbial diversity and altered microbial metabolism. This dysbiosis disrupts immune regulation, gut–brain signaling, and energy balance, increasing susceptibility to obesity and insulin resistance [12].

In 1997 Egger and Swinburn introduced the concept of obesogenic environments to describe the social and environmental conditions that contribute to weight gain at population level [13]. More recently, this perspective has converged with molecular research showing that prior obesity induces lasting transcriptomic and epigenetic reprogramming in adipose tissue. These changes persist after weight loss and predispose individuals to regain weight, a phenomenon described as obesogenic memory [14].

In biomedical research, obesogenic memory refers to the persistence of physiological adaptations induced by prior obesity, which predispose individuals to weight regain even after successful weight loss. Initially developed to describe lasting metabolic dysfunction in adipose tissue, the concept has expanded to include transcriptomic and epigenetic changes, as well as alterations in central appetite and energy regulation. Crucially, these physiological inscriptions are likely to persist even when weight loss is achieved pharmacologically, including through GLP-1RA treatment.

Despite its explanatory potential, obesogenic memory has remained largely confined to individual physiology, focusing on mechanisms such as adipocyte hyperplasia, metabolic set points, and appetite regulation [15]. As a result, it offers limited conceptual leverage for understanding how obesogenic processes are socially patterned, collectively reproduced, and historically sustained. We propose extending obesogenic memory beyond individual biology to conceptualize it as a multilevel phenomenon encompassing biological and sociocultural processes.

This extension draws on social science theories of memory, which conceptualize remembering as collective, embodied, and materially embedded rather than as an exclusively individual cognitive function. The classic studies on collective memory demonstrate that remembering is structured through social groups, shared practices, and environments, such that individuals remember “through others” [16]. Subsequent theorization emphasizes how memory is sedimented in the body through habitual and performative practices, while it describes forgetting as an active and generative process that enables change and the reconfiguration of identities [17, 18]. Anthropological research further conceptualizes memory as embodied and sensory [19], identifying food practices as a particularly powerful medium through which social memory is formed, reproduced, and contested [20].

More recent materialist approaches conceptualize memory as emerging from dynamic relations between brains, bodies, objects, and infrastructures, assigning analytic agency to material conditions and substances in the production of memory [21, 22]. Dietary practices are then not merely choices but are the outcomes of sociomaterial assemblages that stabilize certain metabolic trajectories over time. Empirical studies of food and memory show how eating practices are reproduced across generations and embedded within familial, collective, and historical contexts [23, 24], complicating behavioral and individualist models of dietary change.

Building on these theoretical foundations, we conducted an in-depth sociological study of people living with obesity to examine how obesogenic memory operates across multiple social domains. We conceptualize obesity not only as a result of transgenerational transmission within families but also as shaped by the metabolic histories of broader collectives, including social and ethnic groups, communities, and nations embedded in specific food production and distribution regimes [25, 26]. Mundane objects and infrastructures—such as food packaging, kitchen technologies, retail environments, and urban layouts—are analyzed as material carriers of obesogenic memory.

Finally, we theorize rescribing obesogenesis as a critical counter-process to remembering. Rescribing refers to the reconfiguration of the sociomaterial patterns that sustain obesogenic memory. Rather than framing change as a matter of individual motivation or knowledge, this perspective foregrounds how modifying material arrangements and infrastructures can enable the unlearning of entrenched metabolic patterns [27, 28]. We hypothesize that conceptualizing obesogenic memory as a dynamic, multilevel biological and sociocultural process provides a robust theoretical framework for reorienting obesity interventions toward collective, systemic, and population-based strategies capable of supporting long-term metabolic health.

## Methods

This article draws on qualitative research conducted within the interdisciplinary project “Technocultures of Extended Metabolism,” led by the Institute of Sociology of the Czech Academy of Sciences in collaboration with the Centre for Research on Nutrition, Metabolism and Diabetes at the Královské Vinohrady University Hospital and the Third Faculty of Medicine of Charles University in Prague. The analysis presented here is based on 49 semi-structured interviews with individuals who self-identify as living with obesity (n=31) and healthcare professionals (n=18). All the participants were Czech. Sampling followed a convenience strategy. Fourteen participants were recruited through an open call published on the social media page of the NGO My Body Is Mine, which organizes educational activities on body shaming and stigma prevention.; the remaining participants were referred by collaborating clinicians. Healthcare professionals were recruited based on publicly available information on their clinical expertise. These clinicians represented a range of specialties involved in obesity care, including internal medicine, diabetology, nutritional therapy, bariatric surgery, and endocrinology.

Interviews were conducted by Varvara Borisova (female), an advanced PhD student in social anthropology at the time of fieldwork, using a semi-structured topic guide. The interviews with healthcare professionals focused on treatment protocols, pharmacological and surgical interventions, and the organization of obesity care within the Czech healthcare system. The interviews with patients addressed treatment experiences, cooking and eating practices, and trajectories of weight loss and weight regain. The duration of the interviews ranged from 46 minutes to 2 hours and 31 minutes. All the interviews were audio recorded and transcribed verbatim in Czech, yielding approximately 1,400 standard pages of text.

The dataset was supplemented with 41 written personal testimonies provided by My Body Is Mine, authored by and with the consent of individuals living with overweight or obesity. In addition, ethnographic fieldnotes were collected during 13 events related to obesity treatment, patient support, and the public health debate, along with methodological reports produced after each interview and observation, totaling approximately 150 standard pages. All the materials cited in this article were translated from Czech into English by the authors.

Data were coded and analyzed abductively by Varvara Borisova and Tereza Stöckelová using Atlas.ti qualitative data analysis software [29]. For this article, analysis focused on codes related to the acquisition, persistence, and transformation of food-related memories, including “familial food habits,” “collective eating,” “national cuisine,” “fatness in the family,” and “changing food habits.” Interpretation was informed by ongoing interdisciplinary collaboration drawing on expertise in biomedicine, public health, and the social studies of medicine.

## Results

This section presents three analytically distinct phases of obesogenic memory identified through abductive analysis: memorizing, remembering, and rescribing. The first phase, memorizing, refers to the assimilation and embodiment of food-related patterns, practices, and sensory preferences. This process involves not only cognitive learning but also habituation to specific flavors and affective associations of tastiness or disgust. The second phase, remembering, captures the persistence and reproduction of these memories through everyday practices of cooking and eating, which are structurally conditioned and embedded in specific material arrangements. The third phase,rescribing, describes processes through which new patterns and routines are acquired, resulting in the creation of new, health-supporting memories.

### Memorizing

A systematic review of longitudinal studies has shown that the family environment plays a crucial role in shaping children’s eating self-regulation [30]. Food parenting practices—defined as “practices that dictate what and how a child eats”—may impair children’s ability to regulate food intake and increase the risk of emotional eating or eating in the absence of hunger [31]. Restrictive feeding practices have also been associated with increased weight gain in children [32]. Parents further shape long-term food preferences by structuring household food environments, modeling eating behaviors, and providing early flavor exposures that generate durable sensory, cognitive, and metabolic responses [33, 34].

Consistent with this literature, most of the narratives traced experiences of obesity and weight loss back to childhood and adolescence, when notions of what counts as “proper,” tasty, or “bad” food were formed and internalized. These notions are transmitted inter- and intragenerationally and shaped by material conditions, family dynamics, and historical context. Across our data, family influence is clustered around three interrelated dimensions: biosocialization into flavors, emotionalization of food, and traumatic postmemories.

Biosocialization into flavors refers to the entangled social and biological formation of food preferences. Ms. Trnka, a woman in her forties who underwent gastric bypass surgery several years ago, recalled that she was “weaned on sausages.” Smoked meats formed the core of her family’s meals, and although she later reduced her consumption of them, the embodied attachment persists:“I managed to cut smoked meats down to a minimum after the bariatric surgery, but that craving is still always there—to have a good, dry piece of salami […] That’s something I’ve really carried with me.”

She retrospectively describes her childhood diet as “terrible” and associates it with later difficulties in weight management, despite years of nutritional education. She recalls limited exposure to fruits and vegetables, shaped less by ignorance than by food and financial scarcity under state socialism and early postsocialism. Affordable, filling, and quick-to-prepare foods structured preferences for processed foods and fast carbohydrates.

These narratives illustrate that flavor preferences emerge through biosocialization—processes conditioned by regional cuisines, ingredient availability, and economic constraints. Extending classical notions of socialization [35], biosocialization emphasizes how early experiences are inscribed in bodies, producing preferences and routines not easily overridden by knowledge or willpower.

Parental feeding practices also shape emotional relationships to food. Mr. Javor, who lost over 50 kg through lifestyle changes and anti-obesity medications, linked his weight gain as an adult to food being used as punishment when he was a child:

“The only punishment in our family was that my parents wouldn’t let me eat. And when I eventually realized that I could eat as much as I wanted, I did.”

Although his family diet was not nutritionally unhealthy, deprivation functioned as a tool of control. Later, overeating became associated with autonomy and relief. Many interlocutors similarly described eating as a coping mechanism, demonstrating how emotionally charged memories shape adult eating behaviors.

Another key mechanism is *postmemory*—the intergenerational transmission of trauma. Ms. Sliva described a household shaped by competing food logics, with a grandmother marked by wartime scarcity and a fat-phobic mother who sought to restrict food consumption. This dynamic exemplifies postmemory as defined by Hirsch and elaborated by Strand [24], where historical trauma persists through everyday practices. In the Czech context, this often takes the form of norms such as finishing everything on one’s plate or avoiding food waste—habits many interlocutors found difficult to unlearn.

Trauma transmission also emerged through body shaming, weight stigma, and internalized fatphobia. Ms. Lipa described a family history marked by eating disorders and anxiety around thinness, which featured as a prominent moral framework. From childhood, she was exposed to strict dietary restrictions and body shaming. She recalls:

“My own eating disorder began as early as kindergarten, when my mother decided that although she had ‘failed’ to stay slim, my sister and I would not. (…) I still remember secretly throwing away my rye bread packed with vegetables and instead finishing my classmate’s white roll with salami—the very food that was forbidden at home.”

Stress linked to these experiences is not only psychological but also contributes to elevated inflammatory markers, including cortisol and C-reactive protein [36], materializing as chronic inflammation. In this sense, stigma-related memories become biologically embedded. Many interlocutors who are now parents described making deliberate efforts to avoid passing on inherited fatphobia to their children.

### Remembering

After obesogenic patterns are memorized, they are actively remembered—that is, re-enacted— through repeated behavioral and emotional patterns that are structurally conditioned beyond the individual. The resulting obesogenic effects are not confined to physiological markers within individual bodies but are continuously produced and reproduced through the social relations, institutions, and material environments in which bodies are embedded [37–39].

Food-related memories formed in childhood play a central role in structuring adult eating practices and affective orientations toward food. For Ms. Hruska, food was closely associated with comfort, care, and emotional security:

“In our family, food was a reward. Food is a kind of comfort…here’s a full plate, go ahead and eat, and don’t worry about anything.”

She recalls that both parents coped with emotions through eating and expressed care through food provision, attachments that continue to shape her relationship to food. Similarly, Ms. Malina describes food-based reward systems as formative but ambivalent:

“At first—until I was about ten or eleven—it was all about rewarding myself with food. But after that, sweets were forbidden, which created a big conflict that I still feel to this day.”

These accounts illustrate obesogenic remembering as an emotionally charged pedagogy of eating, in which food functions as reward, regulation, and moral signal.

Importantly, not all remembered food practices are experienced as obesogenic. Some interlocutors mobilize memories as resources for weight loss. Ms. Jahoda, who grew up in the Carpathian Mountains during the Soviet era, attributes her strong preference for vegetables to conditions of scarcity and subsistence gardening:

“There’s no fruit or vegetable I wouldn’t eat.”

She understands this taste as historically produced and has incorporated it into her own parenting practices. This case shows that remembering can also sustain non-obesogenic trajectories when supported by material and social continuity.

Beyond families, broader collectivities—particularly schools—play a formative role in obesogenic remembering. In Czechia, school nutrition guidelines until 2025 specified food categories but not ingredient quality or processing, enabling compliance through highly processed foods. Largely unchanged for over three decades, these guidelines institutionalized eating practices misaligned with contemporary nutritional science. Schools also shape embodied habits. As Slepičková shows, rules such as enforced speed foster eating practices later remembered and reenacted [40]. Ms. Vrba recalls:

“I was used to ‘throwing’ food into myself…it was the same in school canteens—’hurry up, hurry up’.”

Here, speed and inattentiveness become durable dispositions rather than situational responses.

National cuisines and food-related customs further consolidate obesogenic remembering. Grøn’s discussion of hygge highlights how convivial eating can contribute to obesity while challenging individualized explanations of weight gain [41]. In Czechia, two features are salient: high alcohol consumption—approximately 10 liters of pure ethanol per capita annually—and a cuisine centered on meat, fats, and carbohydrate-heavy side dishes [42]. Although many of the interlocutors said they now avoid these “Czech classics,” they remain culturally unavoidable at celebrations and social gatherings. Obesogenic remembering thus persists through national belonging and shared culinary repertoires, even when individuals attempt to diverge from them.

### Rescribing

If obesogenic remembering describes how eating practices are stabilized over time, rescribing captures the effort to reconfigure these pathways. As Professor Topol noted, successful weight loss requires “forgetting how it was”—a formulation that points to deliberate disengagement from prior food memories. Lifestyle interventions can therefore be understood as practices of rescribing, aimed not merely at changing diets but at reworking social relations, routines, and affective attachments.

Drawing on Connerton’s conceptualization of forgetting as a productive rather than a deficient process, rescribing can be understood as a form of intentional forgetting that enables new forms of embodiment [18]. Mr. Habr’s experience illustrates both the possibilities and limits of this process. Having lost significant weight prior to the Covid-19 pandemic and now restarting treatment with Mounjaro, he emphasizes that obesity in his family is transmitted through shared practices as much as genetics. His parents’ unceasing attempts to provide him with foods that he tries to avoid actively undermines his efforts:

“They think they’re doing it to make me happy, but really, they’re making themselves happy…If you know I don’t eat it, why do you keep buying it?”

For Mr. Habr, maintaining weight loss required not only acquiring new habits but also distancing himself from family-based food relationships that no longer supported his identity. He even contemplated limiting parental visits to protect his rescribed practices. This underscores that rescribing may necessitate partial withdrawal from key sites of obesogenic remembering.

Our clinical interlocutors consistently emphasized that such distancing is often unevenly available. Treatment outcomes, particularly for women, depend heavily on partner and family support. As Dr. Lipa observed, unsupportive partners who tempt patients to abandon dietary changes can critically undermine progress, whereas shared commitment to rescribing increases the likelihood of success. Obesity treatment thus unfolds within relational fields that can either stabilize or destabilize newly formed practices.

Pharmacological interventions—especially novel injectable incretin-based therapies, GLP-1/GLP-1 and GIP receptor agonists—introduce new dynamics into rescribing processes. These drugs not only suppress appetite but, in some cases, alter taste preferences [43]. As Ms. Svestka explains:

“I used to really crave sweets…Now I don’t miss sweets at all.”

However, existing evidence suggests that once medication is discontinued, obesogenic memories often reemerge rapidly [44]. This indicates that pharmacotherapy alone does not erase obesogenic remembering. Instead, it may create a temporal window in which new memories, relationships, and practices can be established. Without such concurrent rescribing, discontinuation risks reactivating entrenched pathways and can lead to weight regain [45]. Sustainable outcomes depend on whether new foodways are socially and materially supported. As Mr. Habr’s case demonstrates individual rescripting can succeed temporarily, but collective remembering—embedded in families, institutions, and national cuisines—often remains intact. Even in the era of GLP-1 therapies, transforming systemic conditions that sustain obesogenic remembering remains essential for long-term metabolic and social health.

## Discussion

Our sociological investigation identified three analytically distinct but empirically overlapping phases of obesogenic memory: memorizing, remembering, and rescribing (Fig. 1). Memorizing refers to the initial inscription of food-related dispositions and practices, typically—but not exclusively—during childhood. Remembering denotes the stabilization and reenactment of these dispositions in everyday life. Rescribing captures efforts to disrupt, overwrite, or forget obesogenic pathways. These phases are not strictly sequential. Individuals may cycle through them repeatedly [46], and different lifestyle domains—diet, physical activity, or sleep—may occupy different phases simultaneously.

**Fig. 1.**
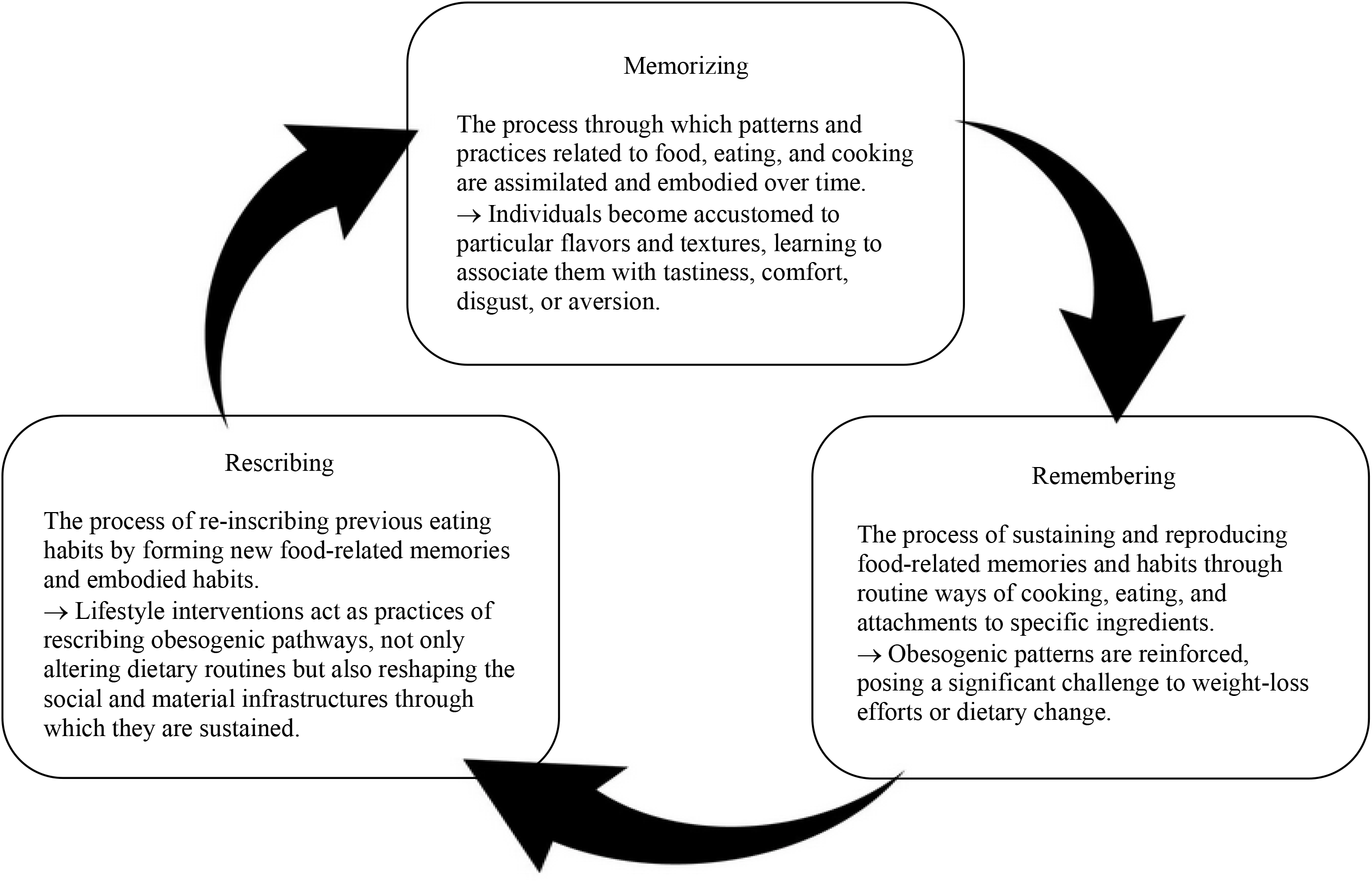
The three-phase cycle of obesogenic memory (schema created by the authors). The memorizing phase describes the initial acquisition and engraving of obesogenic influences. The remembering phase captures the habituation of these influences, contributing to obesity maintenance. The rescribing phase represents the processes through which obesogenic influences may be attenuated or reversed, creating conditions for sustainable health behavior change.

If obesogenic memory is a systemic and collective process rather than merely an individual biological phenomenon, interventions based solely on individual energy balance and lifestyle modification are insufficient. A shift in intervention logic is therefore required: rather than targeting isolated bodies, preventive and therapeutic strategies must engage the collectivities and infrastructures through which food and eating are memorized and reproduced. This reframing shifts the focus from individual behavior change to the biosocial and sociomaterial environments that sustain obesity. Consequently, interventions should extend beyond individual-level strategies to encompass the shared imaginaries, practices, and infrastructures that shape obesogenic risks.

The memorizing phase is anchored in early-life experiences and first encounters with food cultures. Here, individuals learn not only what they like, but what they are expected to like. Interventions at this stage include family-based approaches and early childcare and school settings. Within families, practices such as shared meals, mindful eating, and involving children in food procurement and preparation are central. Cooking skills, in particular, function as a form of embodied resilience. Equally important is access to diverse, minimally processed foods, including fruits and vegetables. The use of fiscal instruments to address food deserts, such as health insurance providers offering reimbursements to incentivize the purchase of healthy food baskets, can act as a structural complement to household-level efforts.

Family-based interventions, however, encounter limits imposed by social inequality. Kindergartens and schools therefore constitute critical sites for interventions during the memorizing phase. School catering should move beyond meeting minimum nutritional requirements and become an active medium for transmitting food culture and health-related values. For many children from disadvantaged backgrounds, the school lunch is the only complete meal of the day. What appears on the plate—and how it is served—teaches children what counts as food, health, and normality. Nutritional education and basic cooking skills should be integrated into school curricula alongside physical education and food culture should be recognized as a core component of health education.

In the remembering phase, obesogenic memory is already stabilized. Interventions here aim to facilitate rescribing through both individualized medical approaches and systematic environmental change. These could include fiscal measures that gradually modify consumer behavior, urban and retail design that increases access to healthy foods, and community-based food infrastructures such as shared tables, outdoor kitchens, and neighborhood dining initiatives. Additional measures include responsible media representations of obesity, public education on the complexity of obesity, and destigmatization campaigns. Several countries have already introduced restrictions on advertising sugar-sweetened beverages, front-of-package labeling for ultra-processed foods, and regulations analogous to those applied to tobacco products [47]. Particular attention must be paid to high-risk populations, including people living in poverty, socially excluded communities, and night-shift workers.

The rescribing phase represents the most explicit attempt to rewrite obesogenic memory. Forgetting, in this sense, entails acquiring new skills and constructing alternative, more resilient memories. At the individual level, induced forgetting is difficult, which helps explain the limited long-term effectiveness of lifestyle interventions. For some individuals, eating regulation is entangled with early-life trauma, making them candidates for cognitive-behavioral or addiction-oriented approaches that address craving, high-risk situations, and emotional regulation.

A major paradigm shift is currently underway with the introduction of incretin-based pharmacotherapies. Although these are typically framed as individually targeted treatments, their rapid spread places them on the threshold of population-level intervention. This raises critical questions about their collective effects. As grocery retailers adjust product offerings in response to growing numbers of GLP-1 users [48], these medications may inadvertently contribute to the rescribing of shared food environments. Whether this moment results in improved metabolic milieus or merely new forms of pharmacological dependency depends on how these shifts are governed.

Incretin-based therapies thus open a window of collective opportunity. If aligned with broader interventions that improve food quality and access and the social relations of eating, they may support a durable rescribing of obesogenic memory, both individual and collective. If used in isolation, however, they risk reinforcing a cycle in which biological suppression substitutes for structural change. Addressing obesity as a biosocial phenomenon therefore requires coordinated interventions across biological, social, and infrastructural levels—interventions capable not only of treating bodies but also of transforming the memories embedded in the worlds they inhabit.

## Acknowledgement

We would like to express our gratitude to the study participants. We also thank Hana Porkertová, Sabina Vassileva, Bodil Just Christensen, and Pernille Andreassen for their feedback on an earlier draft of this article.

## Statements

### Statement of Ethics

This study was performed in accordance with the Declaration of Helsinki. This human study was approved by Research Ethics Board of the Institute of Sociology of the Czech Academy of Sciences (SOÚ-384_3/2024; 1 October 2024) and by the Ethics Committee for Multi-Centric Clinical Trials of the Královské Vinohrady University Hospital (EK-VP/54/2024; 6 November 2024). All adult participants provided written informed consent to participate in this study. Written informed consent was obtained from the individual(s) for publication of the details of their medical case and any accompanying images for the use of interview transcripts in analysis and publication, contingent upon the protection of their anonymity.

### Author Contributions

VB: conceptualization, study design, fieldwork, data analysis, manuscript writing; TS: conceptualization, study design, data analysis, manuscript writing; JG: conceptualization, study design, manuscript writing. All authors have seen and approved the manuscript.

### Data Availability Statement

All data generated during this study were analyzed for the purposes of this article. The data are securely stored on a protected server at the Institute of Sociology, Czech Academy of Sciences.

To protect research participants, whose narratives cannot be fully anonymized due to the qualitative and context-rich nature of the data, the dataset is not publicly available. Upon reasonable request, the original Czech-language excerpts cited in the article may be made available, subject to obtaining additional consent from the respective participants.

